# Different innate and adaptive immune response to SARS-CoV-2 infection of asymptomatic, mild and severe cases

**DOI:** 10.1101/2020.06.22.20137141

**Authors:** Rita Carsetti, Salvatore Zaffina, Eva Piano Mortari, Sara Terreri, Francesco Corrente, Claudia Capponi, Patrizia Palomba, Mattia Mirabella, Simona Cascioli, Paolo Palange, Ilaria Cuccaro, Cinzia Milito, Alimuddin Zumla, Markus Maeurer, Vincenzo Camisa, Maria Rosaria Vinci, Annapaola Santoro, Eleonora Cimini, Luisa Marchioni, Emanuele Nicastri, Fabrizio Palmieri, Chiara Agrati, Giuseppe Ippolito, Ottavia Porzio, Carlo Concato, Andrea Onetti Muda, Massimiliano Raponi, Concetta Quintarelli, Isabella Quinti, Franco Locatelli

**Author notes:** These authors contributed equally to this work (Eva Piano Mortari, Sara Terreri). **Corresponding author**: Dr. Rita Carsetti, B Cell Pathophysiology Unit, Immunology Research Area, Diagnostic Immunology Unit, Department of Laboratories, Bambino Gesù Children Hospital, Viale di San Paolo 15, 00146 Rome, Italy.

## Abstract

SARS-CoV-2 is a novel coronavirus, not encountered before by humans. The wide spectrum of clinical expression of SARS-CoV-2 illness suggests that individual immune responses to SARS-CoV-2 play a crucial role in determining the clinical course after first infection.

Immunological studies have focussed on patients with moderate to severe disease, demonstrating excessive inflammation in tissues and organ damage. In order to understand the basis of the protective immune response in COVID-19, we performed a longitudinal follow-up, flow-cytometric and serological analysis of innate and adaptive immunity in **64** adults with a spectrum of clinical presentations: **28** healthy SARS-CoV-2-negative contacts of COVID-19 cases; **20** asymptomatic SARS-CoV-2-infected cases; **8** patients with Mild COVID-19 disease and **8** cases of Severe COVID-19 disease. Our data show that high frequency of NK cells and early and transient increase of specific IgA, IgM and, to a lower extent, IgG are associated to asymptomatic SARS-CoV-2 infection. By contrast, monocyte expansion and high and persistent levels of IgA and IgG, produced relatively late in the course of the infection, characterize severe disease. Modest increase of monocytes and different kinetics of antibodies are detected in mild COVID-19. The importance of innate NK cells and the short-lived antibody response of asymptomatic individuals and patients with mild disease suggest that only severe COVID-19 may result in protective memory established by the adaptive immune response.

## INTRODUCTION

SARS-CoV-2 is a novel coronavirus, not encountered before by humans. Thus, everyone is susceptible to infection as the SARS-CoV-2 virus rapidly spreads in the current Coronavirus disease 2019 (COVID-19) pandemic. A wide spectrum of clinical expression of SARS-CoV-2 infection occurs, ranging from asymptomatic to mild upper respiratory tract illness, or moderate to severe disease with respiratory distress and multi-organ failure requiring intensive care and organ support (1). This variability of disease severity suggests that the individual immune responses to SARS-CoV-2 play a crucial role in determining the clinical course after first infection. Understanding the pathogenesis of COVID-19 disease requires in-depth study of underlying immune responses (2). This includes the cellular and molecular basis of the successful protective mechanisms and the role of dysregulated and excessive inflammation (3,4). During the 2003 SARS outbreak, the efficacy of the innate immune responses to SARS-CoV-1 appeared to determine the extent of virus load (5) and adaptive immunity played a critical role during the later stages of infection (6).

In COVID-19, lymphopenia is common and correlates with severity of clinical disease similarly to severe influenza and other respiratory viral infections (7–9). Because of lymphopenia, neutrophil-lymphocyte ratio (NLR) and monocyte-lymphocyte ratio (MLR) increase in patients affected by severe COVID-19 (10,11). Lymphopenia is caused by the reduction of both CD4^+^ and CD8^+^ T cells. Surviving T cells are functionally exhausted and reduced T-cell count predicts an unfavourable clinical course (12,13). T cells able to react to SARS-CoV-2 peptides can be demonstrated in healthy individuals, partly because of cross-reactivity with previous infections by other coronaviruses (14) and are expanded in individuals convalescent from COVID-19 (15).

Antibodies to SARS-CoV-2 are produced in large amounts in patients with severe disease, two-three weeks after the occurrence of first symptoms (16). The role of antibodies in viral elimination is supported by the successful use of convalescent plasma in patients with severe COVID-19 (17). Neutralizing antibodies are directed against the Receptor Binding Domain (RBD) or to other regions contained in the S1 subunit of the Spike protein (18–20). Whilst immune responses to novel antigens encountered for the first time, are first dominated by antibodies of IgM isotype, followed by IgG (21,22), the kinetics and protective or deleterious nature of the antibody responses to SARS-CoV-2 remains to be defined. Initials studies suggest that IgG may be produced earlier or at the same time than IgM (16,23,24). A recent study indicated that the IgA response to SARS-CoV-2 may be rapid, strong and persistent (25,26). The observation that the highest antibody levels are found in patients with severe COVID-19 disease led to the suggestion that antibodies to SARS-CoV-2 may be damaging or ineffective rather than protective (27-29), as was reported from very sick patients with Middle East respiratory syndrome (MERS) (30).

In order to identify the immunological features associated to the different clinical forms of SARS-CoV-2 infection, we performed a longitudinal study by standard 7-9 colour flow-cytometry comparing innate and adaptive immune populations of adults with asymptomatic SARS-CoV-2 infection, mild and severe COVID-19 disease and healthy SARS-CoV-2 negative contacts. We also measured levels and kinetics of IgG, IgA and IgM anti SARS-CoV-2 antibodies in the serum.

## MATERIALS AND METHODS

### Study design

Prospective, parallel-group, design. Patients were enrolled in in-patient and out-patient settings, if they agreed to participate and fulfilled the inclusion/exclusion criteria. Sixty-four adult patients were enrolled in the study (**Supplementary Table S1**). At enrollment, after the protocol procedures (including medical history, physical examination, laboratory examination) participants were assigned to the study group: a) Contacts of SARS-CoV-2 confirmed cases who were negative by qPCR and were included as control group (28 patients); b) Asymptomatic cases (20 patients) tested positive for viral RNA and had no symptoms. Asymptomatic patients were quarantined and monitored for 14 days, and quarantine ended when two consecutive nasopharyngeal swabs showed negative results; c) Mild COVID-19 disease (8 patients), defined by positive SARS-CoV-2 nasopharyngeal swab qPCR test, with symptoms such as fever, myalgia and fatigue without obvious chest HRCT findings for COVID-19, did not require hospitalization; d) Severe COVID-19 disease (8 patients) were hospitalized for respiratory disease with bilateral lung infiltrates at HRCT highly suggestive of COVID-19 interstitial pneumonia, and P/F ≥ 300 mmHg (defined as the ratio of arterial oxygen tension [PaO2] to inspiratory oxygen fraction [FiO2]).

Contacts, asymptomatic individuals and patients with mild disease were Health Care Workers (HCW) of the Bambino Gesù Children Hospital. We included in the study all HCWs that had a positive swab in the period between March 15 and May 31, 2020 and their contacts (who had a negative swab). Blood and serum samples were collected at weekly intervals since diagnosis. Severe cases were patients from the Pulmonary division of the Department of Public Health and infectious diseases, Policlinico Umberto I Hospital, Rome, Italy.

Additional 77 patients with severe COVID-19 were recruited from the INMI, Lazzaro Spallanzani.

### Ethical approval

Ethical approval was obtained from the Medical Research and Ethics Committee at Sapienza, University of Rome and from the Ethics Committee at INMI, Lazzaro Spallanzani. According to the guidelines on Italian observational studies as established by the Italian legislation about the obligatory occupational surveillance and privacy management, HCWs confidentiality was safeguarded and the informed consent was obtained from all the participants. The study was performed in accordance with the Good Clinical Practice guidelines, the International Conference on Harmonization guidelines, and the most recent version of the Declaration of Helsinki.

### Flow-cytometry and antibodies

Four leukocyte profiling panels computing 7- to 9-surface marker antigens for monitoring the major leukocyte subsets as well as characteristics of T-cell, B-cell, monocytes and NK cells subsets were designed (**Supplementary Table S2**). Results of immune-profile of analyzed patients are reported in **Supplementary Table S3-10**. The graphs of the single time points refer to the sample obtained immediately after the first positive nasopharyngeal swab. When available, we also show the results obtained at different time points.

1 ml of total blood (EDTA) was incubated with the lysing solution Pharm Lyse (BD) to lyse red blood cells. Then, cells were divided in four equal aliquots and stained with the appropriate combination of fluorochrome-conjugated antibodies (**Supplementary Table S2**) to identify immune cell subsets according to standard techniques. For the staining of Supplementary Figure 7, heparinized blood of three healthy donors were isolated by Ficoll Paque™ Plus 206 (Amersham Pharmacia Biotech) density-gradient centrifugation. Peripheral blood mononuclear cells (PBMCs) were then stained with antibodies against CD19, CD24, CD27, CD38, IgM, IgG, IgA and IgD (**Supplementary Table S2**). Cells were acquired on a BD FACSLyric™ (BD Biosciences). Data were analyzed with FlowJo ver. 10 (Treestar). Dead cells were excluded from analysis by side/forward scatter gating.

### Serum samples

Included in this study were 169 serum samples obtained from subjects with available clinical records. In particular: fifty-one sera from SARS-CoV-2 negative contacts, sixty-three from SARS-CoV-2 asymptomatic patients, thirty-one from COVID-19 mild patients and fifteen from COVID-19 severe patients. 86 samples from 28 patients were collected at different time points. All sera were kept on ice after collection and then stored at −80°C.

### Serological assays

The Euroimmun Anti-SARS-CoV-2 ELISA IgG and IgA assays (Euroimmun,) were performed on serum samples according to the manufacturer’s instructions. The recommended serum samples dilutions used were 1:100; in samples in which the IgA or IgG quantity was not detectable (overflow), we used 1:1000, 1:3000, 1:6000, 1:25000 dilutions. Values were then normalized for comparison. These ELISA assays provide a semi-quantitative *in vitro* determination of human antibodies of the immunoglobulin classes IgG and IgA against the SARS-CoV-2. The microplate wells are coated with recombinant S1 structural protein. The results were evaluated by calculation of the ratio between the extinction of samples and the extinction of the calibrator. The ratio interpretation was as follows: <0.8 = negative, ≥ 0.8 to < 1.1 = borderline, ≥ 1.1 = positive.

For the detection of IgM anti RBD we developed an in-house ELISA. 96-well plates (Corning) were coated overnight with 1μg/mL of purified SARS-CoV-2 RBD protein (Sino Biological). After washing with PBS/0.05% Tween and blocking with PBS/1% BSA, plates were incubated for 1h at 37°C with diluted sera. Serum samples were measured at 1:100 dilutions. After washing, plates were incubated for 1h at 37°C with peroxidase-conjugated goat anti-human IgM antibody (Jackons ImmunoResearch Laboratories). The assay was developed with *o*-phenylen-diamine tablets (Sigma-Aldrich) as a chromogen substrate. Absorbance at 450 nm was measured, and IgM concentrations were calculated by interpolation from the standard curve based on serial dilutions of monoclonal human IgM antibody against SARS-CoV-2 Spike-RBD (Invivogen). Due to the unavailability of a human IgM antibody against SARS-CoV-2 S1 to be used as standard we were unable to quantify the level of anti-S1 IgM. We could, however compare the OD measured in plates coated with either RBD or S1 (data not shown).

### Statistical analysis

For the comparison of more than two independent groups, the non-parametric Kruskal-Wallis test was used and, if significant, pairwise comparisons were evaluated by the Mann-Whitney *U*-test. P values less than 0.05 were considered statistically significant.

## RESULTS

### Clinical Characteristics

SARS-CoV-2 asymptomatic patients (20 patients, M/F 4/16, mean age, 40.4 years, range 27-64) and SARS-CoV-2-negative contacts (28 patients, M/F 8/20, mean age, 40.8 years, range 27-68) had comparable demographic characteristics. Severe adult COVID-19 patients (8 patients, M/F 6/2, mean age, 65 years, range 30-90) and mild-symptoms adult COVID-19 patients (8 patients, M/F 5/3, mean age, 55.2 years, range 48-64) were older than asymptomatic patients and controls (**Supplementary Table S1**). During the study period, disease activity was regularly assessed, and COVID-19 patients continued their therapies according to the standard of care. Four out of eight severe cases were treated with anti-IL-6R monoclonal antibody (tocilizumab). All hospitalized COVID-19 patients were discharged and none died.

### Innate immunity

The PBMCs of patients with asymptomatic infection, mild and severe disease and their healthy contacts were compared. We correlated the immunological findings with the clinical course and studied the dynamic changes of cells of innate and adaptive immune response in time by analysing blood samples obtained at weekly intervals beginning on the first or second week after diagnosis.

By flow-cytometry performed on the first sample obtained after diagnosis, we confirmed the increase of MLR in advanced COVID-19 cases, when T cells, normally representing the major lymphocyte population in the peripheral blood, are reduced (31) (**Figure 1A and B**). Previous studies indicated that neutrophils and macrophages infiltrate the lungs and are expanded in the peripheral blood of Intensive Care Unit (ICU)-admitted patients (32,33). The increase of circulating neutrophils and monocytes, along with lymphopenia, explains why the NLR and MLR are significantly higher in patients with severe COVID-19 disease (10).

**Figure 1:**
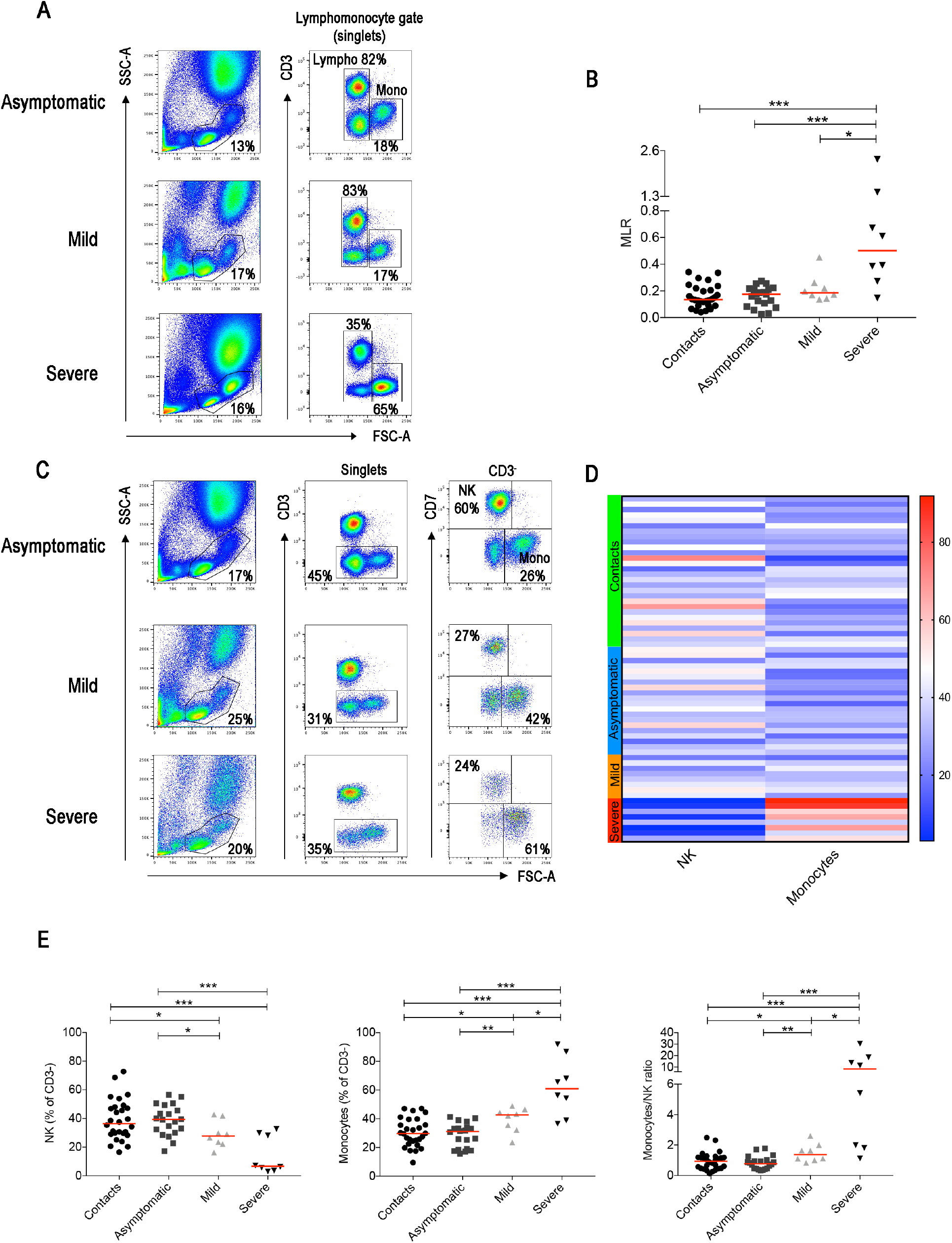
(**A**) Flow-cytometry analysis of the monocyte to lymphocyte ratio (MLR) in the blood of three representative patients with asymptomatic, mild and severe disease. The lympho-monocyte gate was designed based on physical characteristics (FSC-A *vs* SSC-A). Lymphocytes were gated as FSC-A^low^ and CD3^+^ or CD3^-^ and monocytes as CD3^-^ FSC-A^high^. (**B**) Scatter plot depicts the MLR in the sixty-four adult patients enrolled in the study (contacts *n=28*; asymptomatic *n=20*; mild n=8; severe *n=8*). (**C**) Gating strategy used to identify natural killer (NK) and monocytes inside the CD3^-^ cells in three representative patients with asymptomatic, mild and severe disease. NK cells were defined as CD3^-^CD7^+^FSC-A^low^ and monocytes as CD3^-^CD7^-^FSC-A^high^. (**D**) Heatmap shows percentages of NK and monocytes in contacts (indicated by the light green bar), asymptomatic (blue), mild (orange) and severe (red) patients. Percentages are represented by the different expression of red, blue and white as indicated in the color code. (**E**) Plots indicate the frequency of NK, monocytes and the monocytes/NK ratio (MNKR) in our patients. (**B and E**) Midlines indicate median. Statistical significances were determined using unpaired, two-tailed Mann-Whitney *U*-tests. *p≤0.05, **p<0.01, ***p<0.001.

T-cell frequencies are preserved in asymptomatic individuals and in patients with mild disease. Thus, in order to investigate whether other lymphocyte populations than T cells change in asymptomatic and mild disease, we excluded T cells from the analysis (**Figure 1C and Supplementary Figure S1**). The CD3^-^ gate, besides B cells discussed below, includes monocytes that can be distinguished by their larger size measured by the high FCS, and NK cells. NK cells express the markers CD7 (**Figure 1C**) and CD56 (**Supplementary Figure S1**).

We found that NK cells were reduced and monocytes increased in patients with severe COVID-19 (**Figure 1C and D**). Significant reduction of the frequency of NK cells and increase of monocytes were also observed in the group of patients with mild disease (**Figure 1C-E**). We calculated the Monocyte to NK ratio (MNKR), which was <1 in asymptomatic individuals, > 1 in patients with mild disease and even higher in severe cases (**Figure 1E**).

These results were confirmed when we analysed all the samples collected at different time points, as the MNKR remained stable throughout the follow up in asymptomatic and mild disease groups (**Figure 2A**), because each individual maintained his typical NK and Monocyte frequency throughout the time of follow-up (**Figure 2B**). Thus, the different relative frequencies of monocytes and NK observed in asymptomatic, mild disease and severe cases were not incidental findings observed in a particular moment of the infection, but rather characteristics of the clinical course of the response of the individual immune system to SARS-CoV-2 (**Figure 2B**). We confirmed the importance of the frequency of NK cells by the analysis of another independent cohort of 77 patients hospitalized because of severe COVID-19. Cases who did not need ICU treatment had a significantly higher number of NK cells (CD56^+^ cells calculated in CD3^-^ lympho-monocyte gate) than ICU patients (**Figure 2C**). In addition, the percentage of NK cells was low in patients with fatal COVID-19, whereas it increased in those individuals who recovered from severe disease (**Figure 2D**). These results are corroborated by the observation that ICU patients had lower perforin^+^ NK cells compared to non-ICU patients (34).

**Figure 2:**
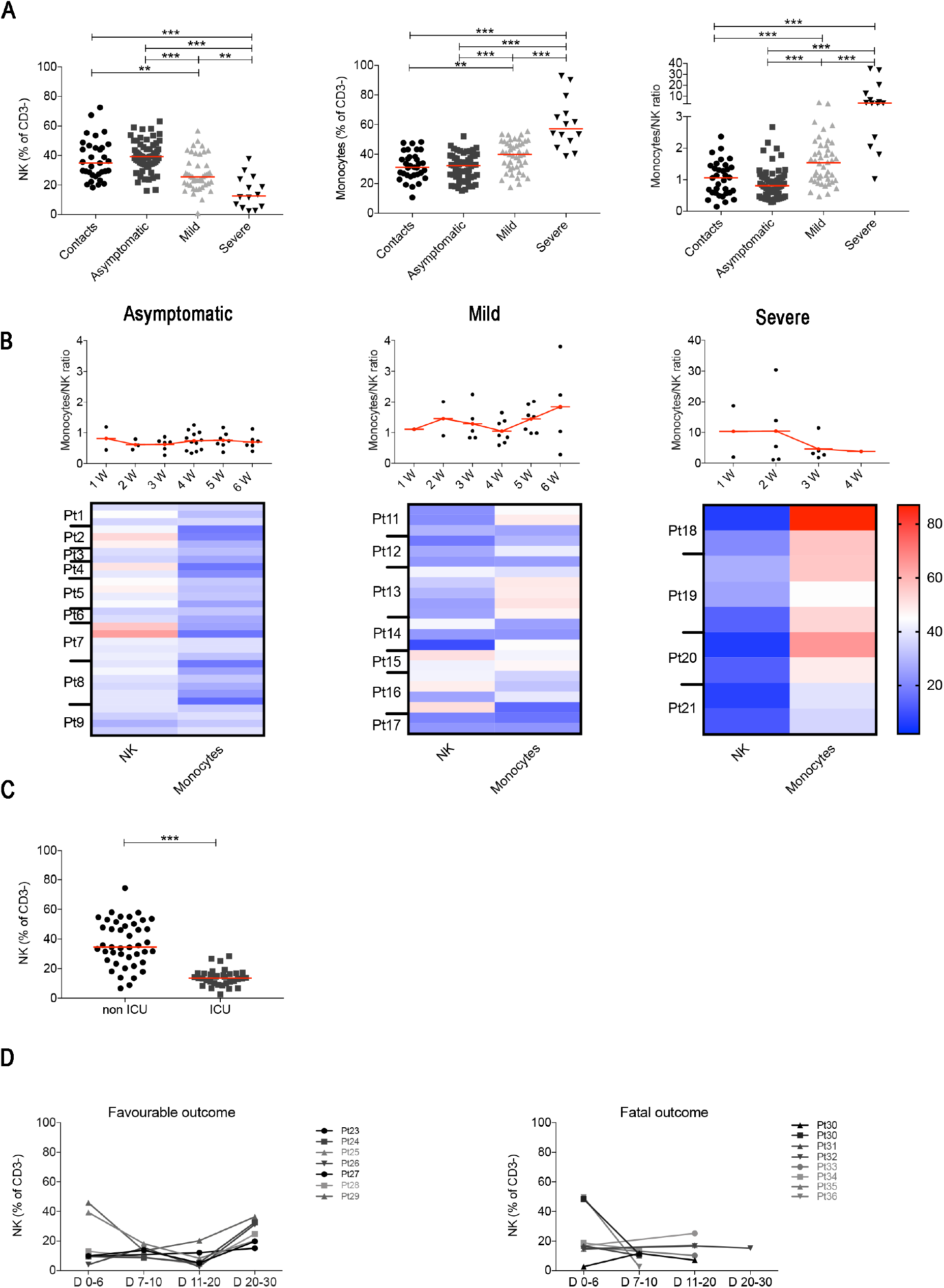
(**A**) Plots indicate the frequency of NK, monocytes and MNKR ratio in all analyzed samples collected at different time points. (**B**) Graphs depict the kinetics of the MNKR during the first 6 weeks of disease (midlines indicate mean) in all patients samples. Data referring to severe patients has a different scale due to the high value of MNKR. Heatmaps show the percentage of NK and monocytes in patients who had samples collected at different time points during the first 6 weeks. In the heatmap percentages are represented by the different expression of red, blue and white as indicated in the color code. (**C**) Plot shows the percentage of NK cells in non-ICU (*n=43*) and ICU (*n=34*) patients. (**D**) Graphs show the kinetics over time of NK cells percentage in patients with favorable and fatal outcome. (**A** and **C**) Midlines indicate median. Statistical significances were determined using unpaired, two-tailed Mann-Whitney *U*-tests. *p≤0.05, **p<0.01, ***p<0.001.

The increase of inflammatory cytokines mostly produced by monocytes plays an important role in determining systemic and local damage in COVID-19. Inflammatory cytokines are produced by intermediate monocytes that expand in the blood of patients with severe infections (35,36). In order to measure the frequency of intermediate monocytes, we used CD16 and CD14 expression to differentiate the three types of CD14^+^ monocytes (**Supplementary Figure S2 and S3A**) found in the peripheral blood, reflecting sequential stages of maturation and distinct functions (37,38). CD14^+^CD16^-^ classical monocytes are the precursors of the other types and play an important role in the response to pathogens (39). CD14^-^CD16^+^ non-classical monocytes contribute to the resolution of inflammation and maintain vascular homeostasis and endothelial integrity (40). Intermediate monocytes express CD14 with variable levels of CD16.

We confirm that, as reported in a recent paper (41) that non-classical monocytes were significantly reduced in patients with severe COVID-19 when compared to SARS-CoV-2 negative contacts, SARS-CoV-2 positive asymptomatic and also mild COVID-19 disease patients (**Supplementary Figure S3B**). Intermediate monocytes tended to increase in the severe cases. As the progression from the classical to intermediate stage is a dynamic step driven by infectious triggers (37), we compared the monocyte phenotype in the same patients at different time points during the course of the disease. Whereas intermediate monocytes were rare in the blood of contacts, asymptomatic and mild disease patients at all time points, transient increases were observed in patients with severe disease (**Supplementary Figure S3C**).

In summary, we found that the MNKR reflects the clinical phenotype of the disease. Contact and asymptomatic patients had either higher representation of NK cells or a similar frequency of NK and monocytes (ratio around 1). The ratio was >1 in patients with mild disease and reached higher values in the severe cases (**Figure 2B**). Thus, the equilibrium between two cell types of the innate immune system may play a role in the control of SARS-CoV-2 infection. Prevalence of NK cells is associated to asymptomatic infection, while increased frequency of monocytes to severe disease.

### Adaptive immunity

T and B cells play key roles in response to viral infections. CD8^+^ T lymphocytes are crucial for the limitation of viral spread through their cytotoxic function. CD4^+^ T cells are indispensable for the expansion of CD8^+^ T cells (42) and the generation of CD8^+^ memory T cells (43,44). In addition, CD4^+^ T cells are necessary for the germinal centre (GC) response and the production of memory B cells (MBCs) and plasma cells (45–47). A recent study reported that 82.1% of the severe COVID-19 cases had low circulating lymphocyte counts because of the reduced frequency of CD3^+^ T cells, both of CD4 and CD8 type (48). In severe COVID-19, not only the number of CD8^+^ T cell declines but also their function is impaired, in association with the increase of pro-inflammatory cytokines (34).

No signs of CD4^+^ and CD8^+^ activation was observed in SARS-CoV-2 positive asymptomatic subjects. Activated, HLA-DR+ CD4 T cells were measurable in patients with both mild and severe disease (**Figure 3A and B**), whereas HLA-DR+ CD8 T cells were only increased in patients with severe COVID-19 (**Figure 3A and B**) in line with previous observations based on transcriptomic analysis (49). These findings were confirmed when we included all samples in the analysis (**Supplementary Figure S4**).

**Figure 3:**
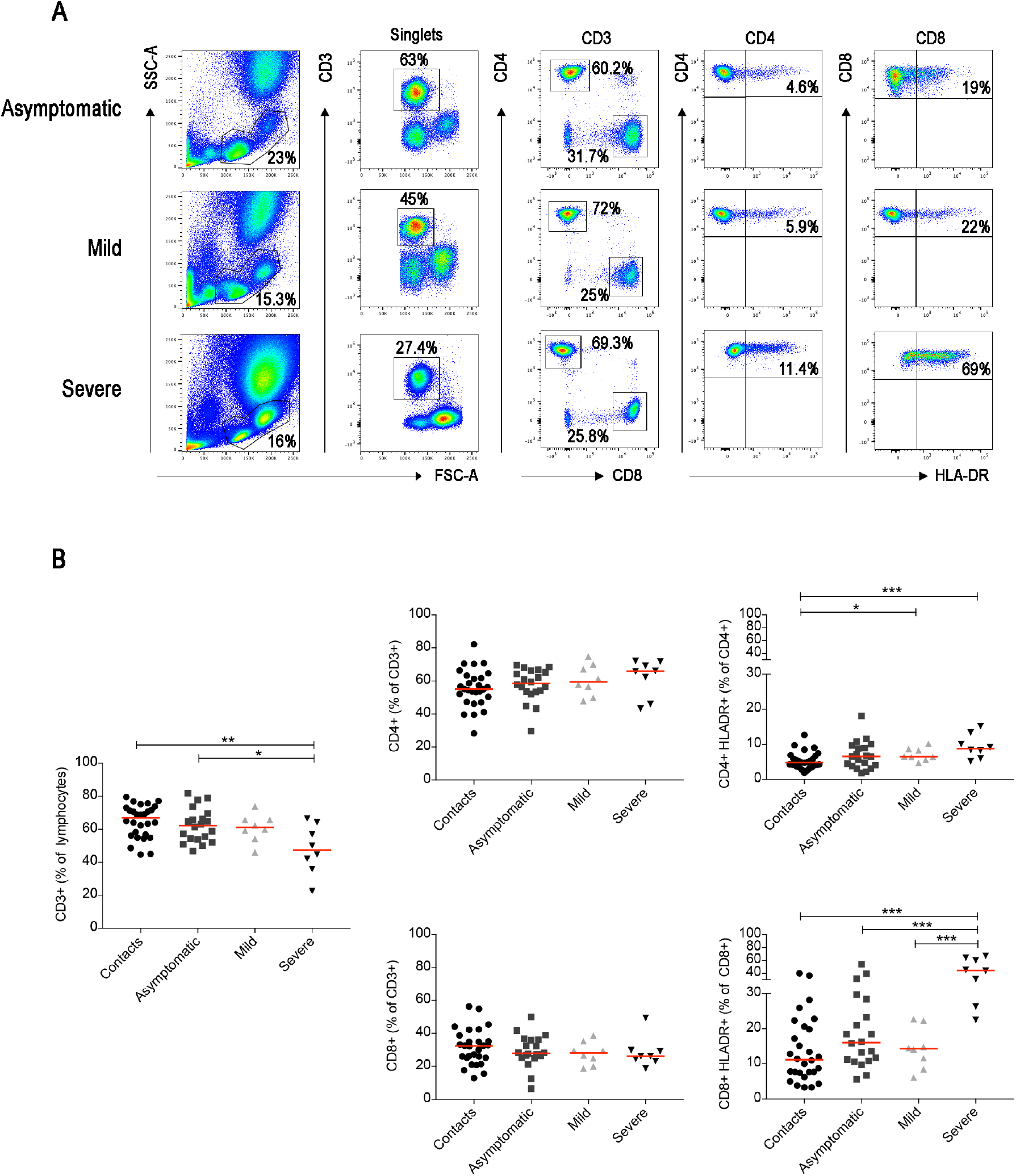
(**A**) FACS plots depict the gating strategy to identify CD3^+^, CD3^+^CD4^+^, CD3^+^CD8^+^ T cells in three representative patients (asymptomatic, mild and severe). HLA-DR expression in CD4^+^ or CD8^+^ T cells is shown in the relative plots. Percentage of HLA-DR^+^ cells are reported in the plots. (**B**) Plots show the percentage of CD3^+^, CD4^+^, CD8^+^ and HLA-DR^+^ T cells as single value. Median is shown as midline. Statistical significances were determined using unpaired, two-tailed Mann-Whitney *U*-tests. *p≤0.05, **p<0.01, ***p<0.001.

In a separate staining, we identified naïve and memory T cells, including central, effector and terminally differentiated (TEMRA) memory T cells (**gating strategy in Supplementary Figure S5**). Also, with this staining we did not found difference between contacts and asymptomatic individuals. In the CD4^+^ population of patients with mild and severe COVID-19, we detected a reduction of recent thymic emigrants (CD45RA^+^CCR7^+^CD31^+^) and a relative increase of *naïve* (CD45RA^+^CCR7^+^CD31^-^) T cells. The frequency of CD4 central, effector memory and TEMRA T cells was not modified by the infection (**Figure 4 and Supplementary Figure S6**). CD8^+^ T cell distribution was only changed in patients with severe disease. CD38 *naïve* T cells were reduced and, as reported before (50,51) CD8^+^ TEMRA were increased (**Figure 4B and Supplementary Figure S6**).

**Figure 4:**
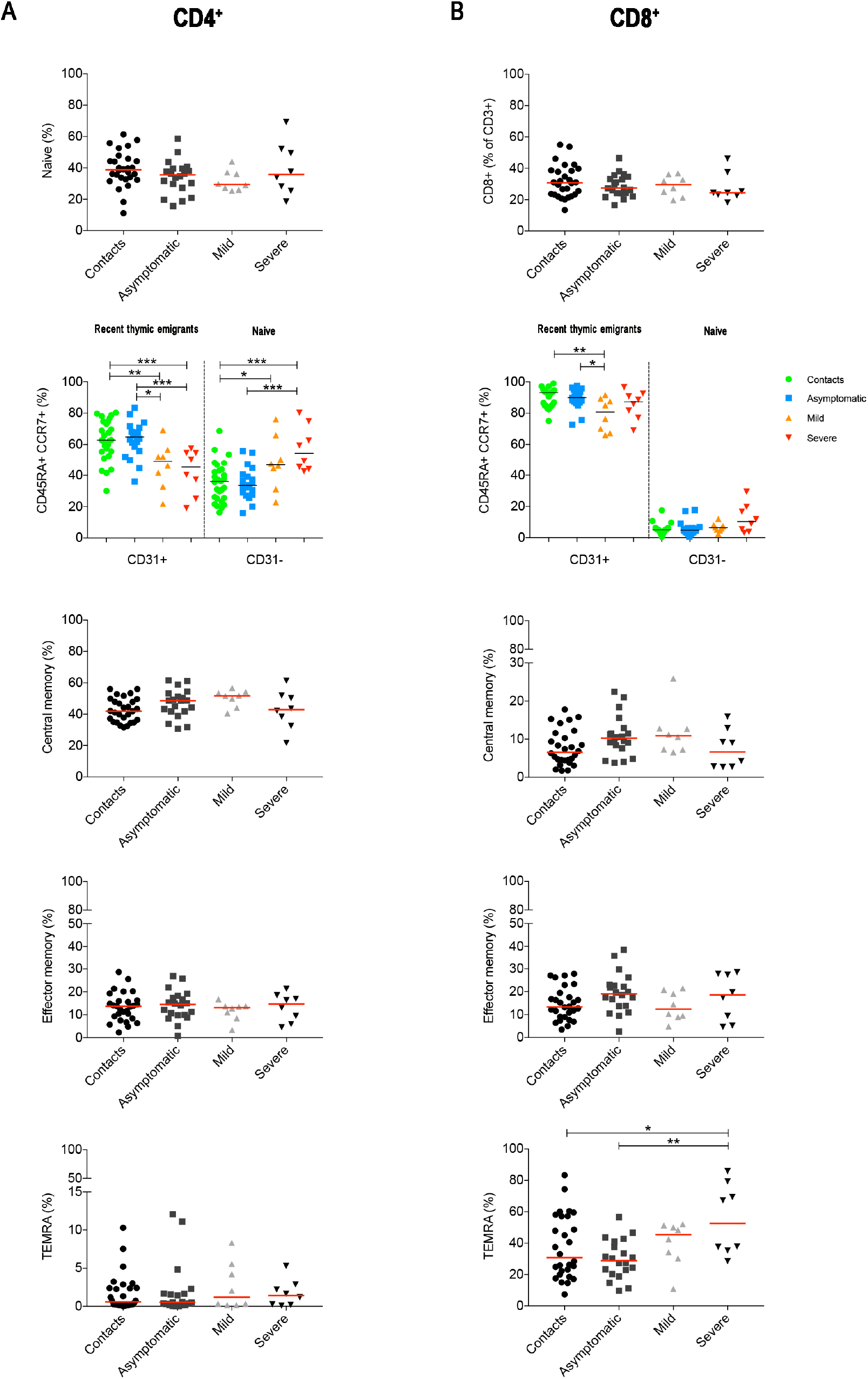
(**A-B**) Results obtained by the analysis of CD4^+^ (**A**) and CD8^+^ T cells (**B**) are separately shown. Graphs show the percentage of *naïve* T cells (CD3^+^CCR7^+^CD45RA^+^) that either expressed or lack CD31 (CD31^+^ are recent thymic emigrants and CD31^-^ are revertant T cells). Central memory (CD3^+^CCR7^+^CD45RA^-^), and effector memory (CD3^+^CCR7^-^CD45RA^-^) T cells and TEMRA (CD3^+^CCR7^-^CD45RA^+^) T cells. Midlines indicate median. Statistical significances were determined using unpaired, two-tailed Mann-Whitney *U*-tests. *p≤0.05, **p<0.01, ***p<0.001.

Our data indicate that the circulating pool of T cells is not altered in the SARS-CoV-2 asymptomatic infection. Expression of HLA-DR on CD4^+^ T cells, reflects their activation in both mild and severe cases. Activation of CD8^+^ T cells could be only demonstrated in the severe cases and was associated to the accumulation of exhausted TEMRA (50,51).

We identified the different B-cell populations in the peripheral blood by staining with a combination of antibodies able to distinguish transitional, *naïve*, memory, atypical MBCs and plasmablasts (PBs) (**Figure 5A**). In the CD27^+^ MBCs population, we separately analysed IgM^+^ and switched MBCs. The latter include IgG^+^ MBCs and IgG^-^ MBCs. Most of the IgG^-^ B cells correspond to IgA-expressing memory B and in minimal part to MBCs without detectable surface immunoglobulin (**Supplementary Figure S7**). The most significant findings were the reduction of total B cells and the increase of PBs in the severe cases (**Figure 5B**), as reported in other studies (52–54). Among MBCs, we found an increase of IgM^+^ and a reduction of switched MBCs in asymptomatic and mild cases. In patients with severe disease, in contrast, we observed an increase of switched MBCs negative for IgG and mostly expressing IgA. All the findings were confirmed by the cumulative analysis of all samples (**Supplementary Figure S8**).

**Figure 5:**
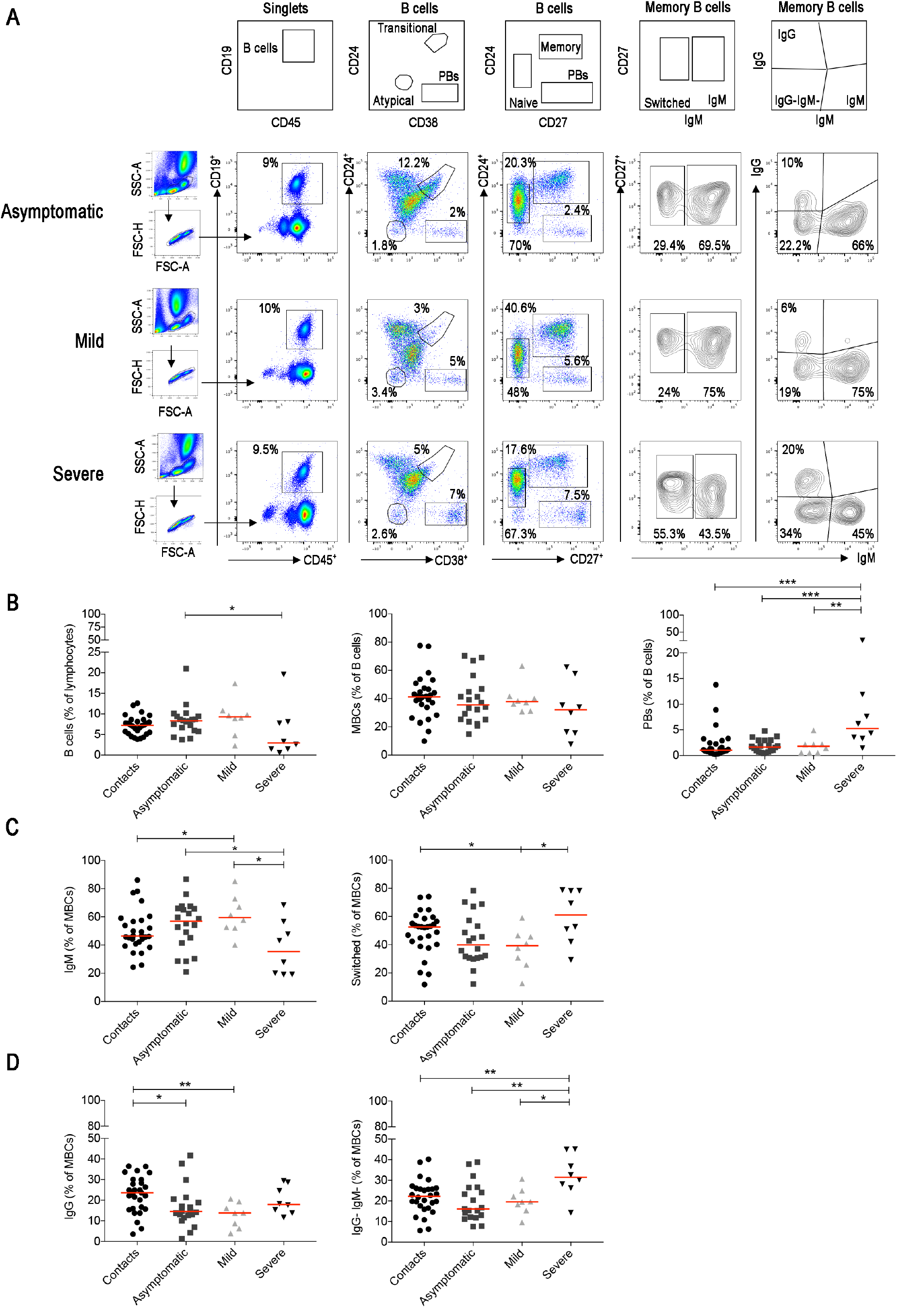
(**A**) Viable lymphocytes were gated and then selected as CD19^+^ B cells in three representative patients with asymptomatic, mild and severe disease. The identification of the different B cell populations is shown in the empty plots of the upper line. We identified transitional (CD24^+^CD38^++^), naïve (CD24^+^CD27^-^), memory (CD24^+^CD27^+^), atypical MBCs (CD24^-^CD38^-^) and plasmablasts (CD24^-^CD27^++^CD38^++^). In the CD27+ memory B-cell population based on IgM expression, we show IgM and switched (IgM^-^) MBCs. MBCs were also gated as IgM^+^, IgG^+^ and IgG^-^ IgM^-^ MBCs. (**B**) Plots indicate the percentage of B cells, MBCs and plasmablasts. In **C** the frequencies of IgM and switched MBCs are shown. In panel **D**) we show the frequency of IgG^+^ and IgG^-^IgM^-^MBCs. Midlines indicate median. Statistical significances were determined using unpaired, two-tailed Mann-Whitney *U*-tests. *p≤0.05, **p<0.01, ***p<0.001.

### Antibodies

B cells fight viruses by producing antibodies when they differentiate into circulating PBs or tissue-resident plasma cells (55). The final stages of differentiation can be reached by B cells in the GCs, where either *naïve* or IgM^+^ MBCs (56,57) acquire somatic mutations and are selected for their increased affinity to the stimulating antigen (46). T- and GC-independent antibody production is efficiently and rapidly triggered by TLR-mediated stimulation of MBCs (58,59).

IgG and IgA antibodies directed against the S1 domain of the SARS-CoV-2 Spike protein were measured in the entire study cohort. We also measured the concentrations of IgM specific for the SARS-CoV-2 RBD.

Both COVID-19 patients and SARS-CoV-2 positive asymptomatic individuals produced specific antibodies, with higher levels of IgG and IgA being detected in the serum of patients with severe disease (**Figure 6A**) as reported before (18,26,28,50).

**Figure 6:**
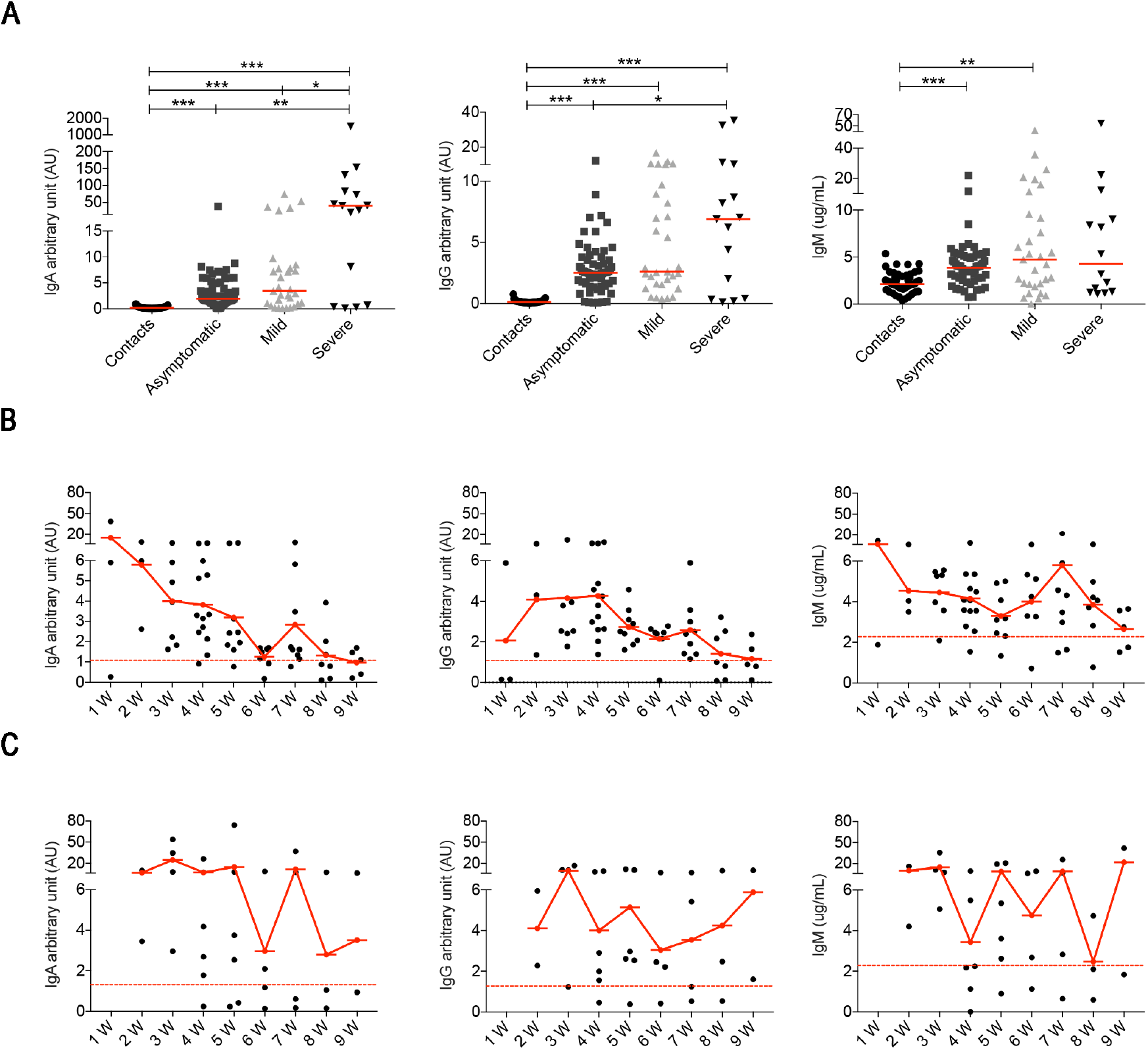
(**A**) Arbitrary units (AU) of IgG and IgA specific for the S1 domain of the SARS-CoV-2 Spike protein and concentration of RBD specific IgM were detected by ELISA at different time points. Data relative to all samples are shown. Midlines indicate median. Statistical significances were determined using unpaired, two-tailed Mann-Whitney *U*-tests. *p≤0.05, **p<0.01, ***p<0.001. (**B-C**) Graphs show the levels of IgA, IgG and IgM during the course of the disease in asymptomatic (**B**) and mild (**C**) patients. Time is indicated in weeks starting from the first positive nasopharyngeal swab. Dashed line indicates detection threshold.

We show in detail the kinetics of the antibody response of asymptomatic and mild disease individuals initiating from the earliest sample obtained after the first positive nasopharyngeal swab (**Figure 6B and C**).

We found that asymptomatic patients secrete specific IgA and IgM early after diagnosis. Levels of IgA and IgG rapidly decline and become undetectable after 8 weeks. As for IgA and IgG there is an established threshold of positivity (red line), we could establish that individuals lacking IgG and IgA at week 1, later produced antibodies that then declined weeks after the first positive PCR. There is no established threshold for anti-RBD IgM, but we found that the median IgM concentration in 54 healthy contacts was 2.1 μg/ml (range 0.3-5.4). We found that IgM levels remained stably over the mean value of contacts in most asymptomatic individuals (**Figure 6B**).

In Figure 6C we show the antibody response of mild disease patients. In this group values below the threshold were found in two individuals, throughout the course of disease. Pt14, an otherwise healthy 48 year-old HCW, never produced IgG, IgA or IgM (5 samples were evaluated). She had neurological symptoms and a positive nasopharyngeal swab PCR for 6 weeks. Pt12 (54-year-old),with respiratory symptoms and aPCR that remained positive for 8 weeks, had very low IgG, IgA and IgM levels fluctuating around the detection threshold. In the rest of the mild disease patients, IgA, IgG and IgM abs increased later than in asymptomatic individuals but remained over the threshold for 9 weeks. In summary, we observed that severe patients showed a strong antibody response in line with other observations (18,26,28,50,60). Levels of IgA and IgG in asymptomatic individuals returned to levels comparable to those measured in healthy contacts 6-8 weeks from the first positive swab. Antibodies did not decline rapidly in mild disease patients.

## DISCUSSION

The first response to a novel virus is typically characterized by the cooperation between NK cells and natural antibodies, key components of innate immune system (61–65). Since the adaptive response requires time to build up after first encounter with an unknown pathogen, NK cells and natural antibodies contain the infection, whilst adaptive immune responses develop and generate highly-specific memory T and B cells that will clear the virus and prevent re-infection (22).

We still do not know whether the infection with SARS-CoV-2 triggers this typical response. We do know that individual immune responses play an important role in determining the clinical course of SARS-CoV-2 infection.

In order to understand the basis of the immune response in COVID-19, we performed a global analysis of innate and adaptive immunity in patients selected across the spectrum of disease severity, ranging from SARS-CoV-2 positive asymptomatic individuals to patients with mild and severe COVID-19. We used standard flow-cytometry and serology with the aim of finding an easy-to-use tool for the clinics.

Our data show that the balance between NK cells and monocytes is a sensitive indicator of the individual reaction to the virus and is related to the clinical course of the disease. We calculated the ratio between the frequency of peripheral blood monocyte and NK cells (MNKR) and found that each individual included in our study maintained his typical MNKR throughout the time of follow-up. The MNKR is below 1 in contacts and asymptomatic individuals and increases when monocytes expand, and NK cells are reduced. This phenomenon occurs in mild COVID-19, when the frequency of NK cell slightly declines and that of monocytes increases and is more and is more evident in patients with severe disease, where the loss of NK cells is associated to the expansion of monocytes. Monocytes secrete inflammatory cytokines causing local and systemic damage (66–68). These alterations are reminiscent of Hemophagocytic Lymphohistiocytosis (HLH), a condition often related to mutations of genes governing the cytotoxic lymphocyte machinery indispensable for the function of NK cells. In HLH, chronic expansion and activation of monocytes causes the life-threatening condition known as cytokine storm (69–72). A similar “storm” is also responsible for the dramatic evolution of severe COVID-19. In these cases, therapeutic strategies aiming at controlling excessive pro-inflammatory cytokine levels have been successfully used (73,74).

In most viral infection the production of type 1 Interferons (IFN-I) promotes NK cell expansion (75) and has a direct anti-viral effects (76). It has been demonstrated that antiviral IFN-I and III are not significantly induced by SARS-CoV-2 infection of respiratory epithelial cells, whereas a chemokines signature is established (77). As a consequence, NK cells survival and function are not supported, but monocytes are attracted to the site of infection. IFN beta-1 was administered as early treatment together with a triple combination of anti-viral drugs in a recently published multicenter, open randomized trial. The therapy was effective in suppressing the shedding of SARS-CoV-2 by acting on virus replication and innate immunity (78). In agreement with our hypothesis it has been demonstrated that inborn error of type I IFN immunity and neutralization of type I IFN function by autoantibodies are associated to the most severe forms of COVID-19 (79,80).

It has been reported that antibodies are produced late in hospitalized patients with COVID-19: IgG increased after three weeks and IgM antibodies were transiently detected often later than IgG (16,24). Earlier antibody production has been shown more recently (81,82).

We measured the specific response to the S1 domain of the SARS-CoV-2 spike protein (IgG and IgA) and to the RBD (IgM) in the serum of all patients and controls in our study at different time points. We confirm that the highest levels of IgG and also IgA are produced by patients with severe disease. Our most interesting observation is the different kinetics of response in asymptomatic and mild disease forms of infection. The early and transient IgM, IgA and IgG response distinguish asymptomatic individuals from mild-disease patients, who have a slower, but more persistent antibody production.

In asymptomatic individuals, the early burst of IgA may rapidly and effectively eliminate the virus in the respiratory mucosa and prevent the development of a full adaptive immune reaction. The slightly slower IgG and IgA production that persist in time suggests that the adaptive immune response is triggered in mild disease and may be able to generate immunological memory. A long and severe disease fully activates the adaptive immune response and is associated with the production of anti-SARS-CoV-2 antibodies, PBs and memory B cells (83). Further studies are necessary to establish whether specific memory persist and for how long after asymptomatic and mild disease.

The particular antigen-specific IgA/IgG profile associated with clinical outcome may reflect TGFbeta production induced by coronavirus-species (84,85). Augmented viral load may increase TGFbeta production, that, if locally secreted in the lung, facilitates neutrophil attraction and specifically induces the isotype switch to IgA (86), a situation that prompted the suggestion of anti-TGFbeta directed immunotherapies (87,88).

Innate MBCs are increased in asymptomatic and mild disease. Innate MBCs produce natural antibodies in response to TLR stimulation (58,89) but are also able to enter the GC where they remodel their antibodies to increase their affinity (56,90). IgM^+^ MBCs are the precursors of most IgA^+^ and IgG^+^ switched MBCs (90) and give rise to IgA^+^ plasma cells at mucosal site (91). ‘Natural antibodies’, produced by innate MBCs, are antibodies that have a protective role in the early phases of the response independently of any previous encounter with antigen (89,92,93). These antibodies, not yet been shaped by antigenic selection, carry few somatic mutations (56) and have broad reactivity (94). We recently suggested that natural antibodies might explain why most pediatric cases with laboratory-confirmed SARS-CoV-2 infection have either no or mild symptoms and recover within 1–2 weeks (95). Cross-reactive antibodies found in children and adult never exposed to SARS-CoV-2 (96) may correspond to natural antibodies. We speculate that the early IgA burst of asymptomatic individuals may derive from the rapid activation of pre-existing innate or cross-reactive IgM^+^ MBCs that switched to IgA in the respiratory mucosa (25), as suggested also by the demonstration that moderate levels of IgM and IgA cross-reactive to SARS-CoV-2 are present in the blood of healthy individuals never exposed to the infection (96). In addition, neutralizing IgG MBCs isolated from COVID-19 patients may have none or very few somatic mutations (97) thus suggesting that the pre-immune or innate MBC repertoire may contain SARS-CoV-2 specificities (82). In patients with severe COVID-19, IgM+ MBCs are reduced and switched MBCs are increased. The increase of switched MBCs may reflect the immune reaction in the lymphoid tissue associated to the respiratory tree for local protection. Circulating PBs are also increased only in the severe cases in correlation with their higher antibody levels.

Our analysis of circulating T cells shows that SARS-COV-2 infection does not alter the T cell pool in asymptomatic individuals. In mild and severe COVID-19, instead, the increase of activated CD4^+^ T cells reflects the ongoing immune activation. Activated CD4^+^ T cells are indispensable for the effector function during acute viral infections and for the expansion of CD8^+^ T cells (98). In severe cases, also CD8^+^ T cell are activated, as reported in a recent study analyzing the immune response in 76 COVID-19 patients from two independent cohorts (52), and TEMRA accumulate in the blood. A persistent viral antigen stimulation and immune dysregulation may lead to T-cell exhaustion, a state of T-cell dysfunction demonstrated to occur during many chronic infections and cancer (99).

SARS-CoV-2 has evolved in bats, which control the infection through their innate immune system, enriched for NK receptors and different types of INF type I genes (100). Bats also produce antibodies that are highly diverse thanks to a repertoire of VH, DH and JH fragments that is much larger than that found in humans (101,102). Antibodies do not undergo further improvement by introduction of somatic mutation. Thus, constitutive IFN type I secretion and ready-to-use antibodies may control viral infection in bats without the need of adaptive immune responses. For this reason, coronaviruses and other viruses remain endemic in bats, without damaging the host (100). Asymptomatic humans may behave like bats, controlling the infection thanks to NK cells and antibodies. The adaptive immune response is strongest in patients with severe disease, following the extensive tissue damage caused by the uncontrolled inflammatory reaction.

Our data may contribute to monitor the clinical disease. Although many large studies have described the inflammatory reaction in severe disease (69,103), clinically it is indispensable to have prognostic markers early in the course of disease in order to promptly choose appropriate treatments (104). The increase of the ratio between Neutrophils (NLR) or Monocytes and lymphocytes (MLR), mainly caused by the loss of lymphocytes, is an indicator of severe disease (3), but does not change in less severe forms when lymphocytes numbers are still maintained. We propose that the monocyte to NK ratio (MNKR) and the levels of specific IgG, IgA and IgM antibodies in the serum may be more sensible early markers of disease evolution. In particular, low level of antibodies in the first two weeks after diagnosis and increase of the MNKR may indicate patients at risk for increased severity of disease.

Herd immunity to a virus is established when the large majority of the population becomes immune and indirectly protects susceptible individuals. Memory T and B cells and long-lived plasma cells generated by the adaptive immune response are the prerequisite for herd immunity, because specific antibodies in the serum and immediately active memory cells prevent individual re-infection and arrest pathogen transmission. Serum antibodies are a useful correlate of protection and have the function of immediate protection in case of re-exposure. Antibodies, as demonstrated by the administration of convalescent plasma, have a protective function in COVID-19. We still do not know whether B and T memory is established after SARS-CoV-2 infection. The rapid decline of specific antibodies in the serum suggests that long-lived plasma cells are not generated in asymptomatic cases. As we still do not know whether long-lasting protective immunological memory is established after SARS-CoV-2 infection, social distancing and infection control will remain indispensable to limit spread of SARS-CoV-2, until effective vaccines are available.

## Data Availability

no datasets or other repositories

## Author Contributions

R.C, S.Z, I.Q, F.L designed the study and performed data analysis and manuscript preparation; E.P.M and S.T performed data experiments and data analysis and helped with manuscript preparation; F.C, C.C, P.P, M.M, S.C, E C, L.M, E.N, F.P performed data experiments; P.P, I.C, C M, V.C, M.R.V, A.S, O.P, C.C, A.O.M, M.R helped with collection of study samples and clinical information. C.A, G.I, C.Q, A.Z, M.M helped with manuscript preparation. All authors approved the final version of the manuscript.

This manuscript has been released as a pre-print at Medrxiv, [CITATION]

---

**Supplementary Figure S1:**
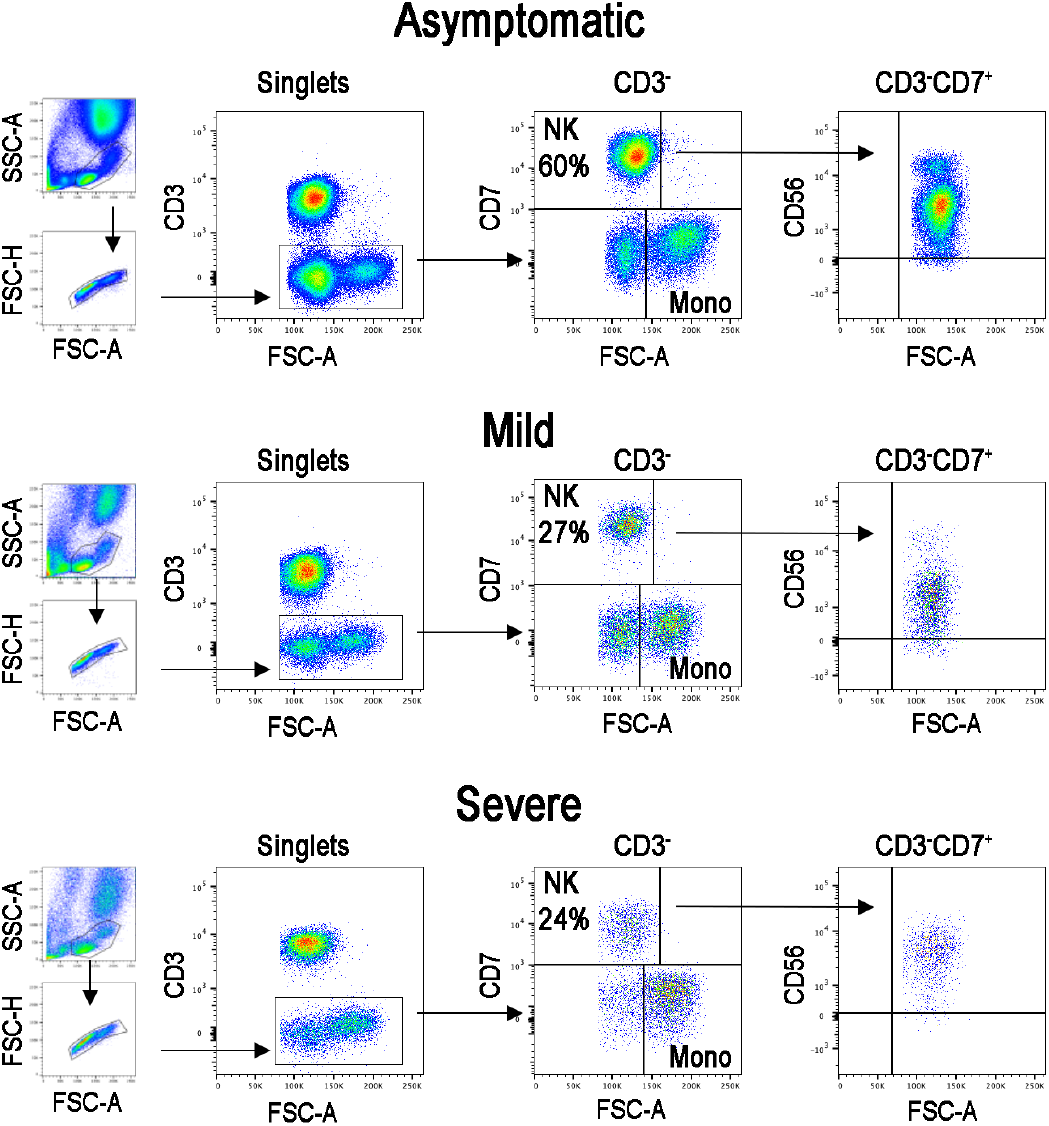
The lympho-monocyte gate was designed based on physical characteristics (FSC-A vs SSC-A). Singlets are identified by FSC-H vs FSC-A parameters. NK were identified as CD3^-^CD7^+^FSC-A^low^ and monocytes as CD3^-^CD7^-^FSC-A^high^. NK cells identified as CD7^+^ in the CD3^-^ gate also express CD56.

**Supplementary Figure S2:**
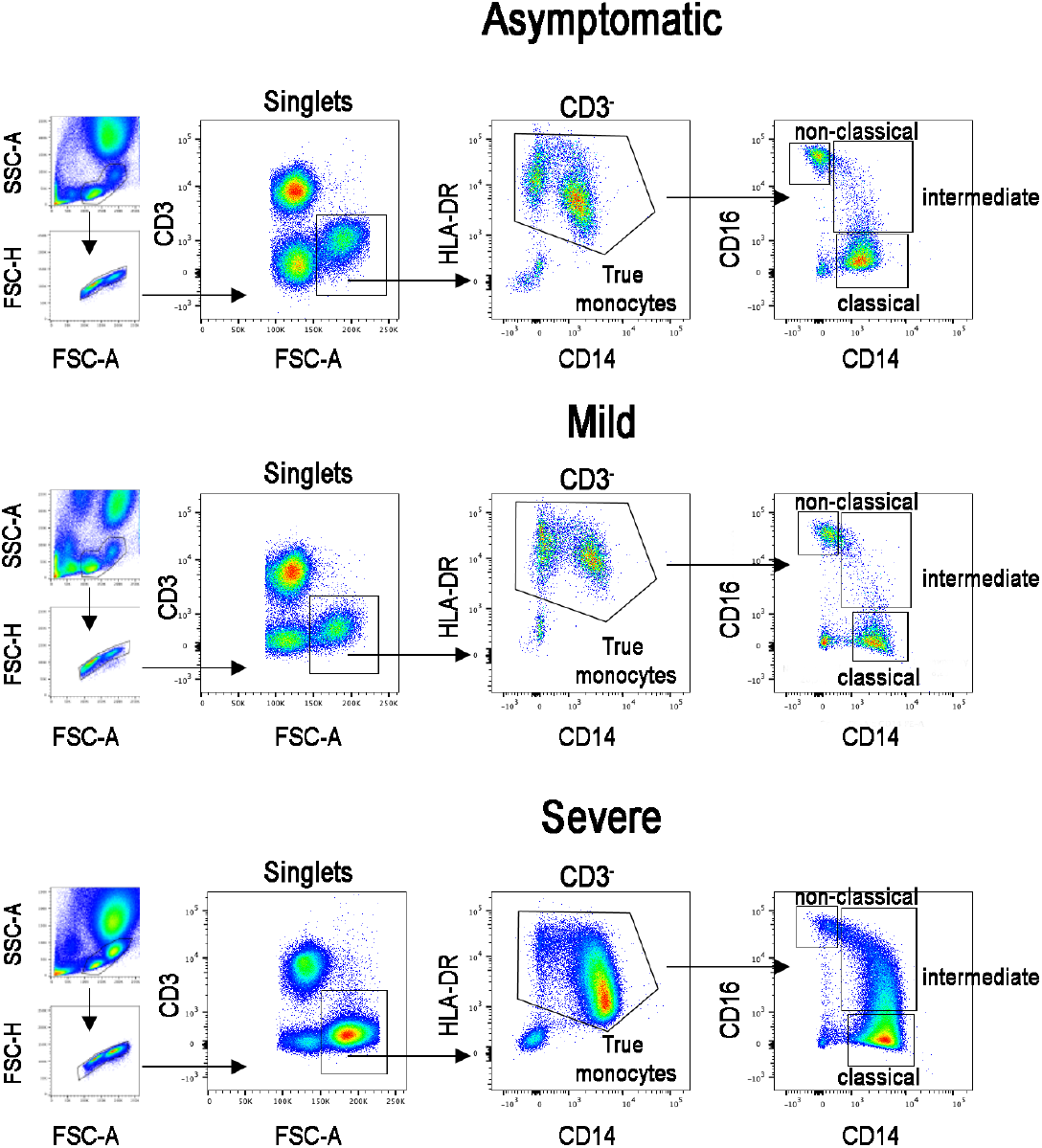
FACS plot show the gating strategy for the identification of monocytes (CD3^-^FSC-A^high^). True monocytes are double positive for HLA-DR and CD14. In the true monocytes gate, we identified the classical (CD14^++^CD16^-^), intermediate (CD14^+^CD16^+^) and non-classical (CD14^+^CD16^++^) populations.

**Supplementary Figure S3:**
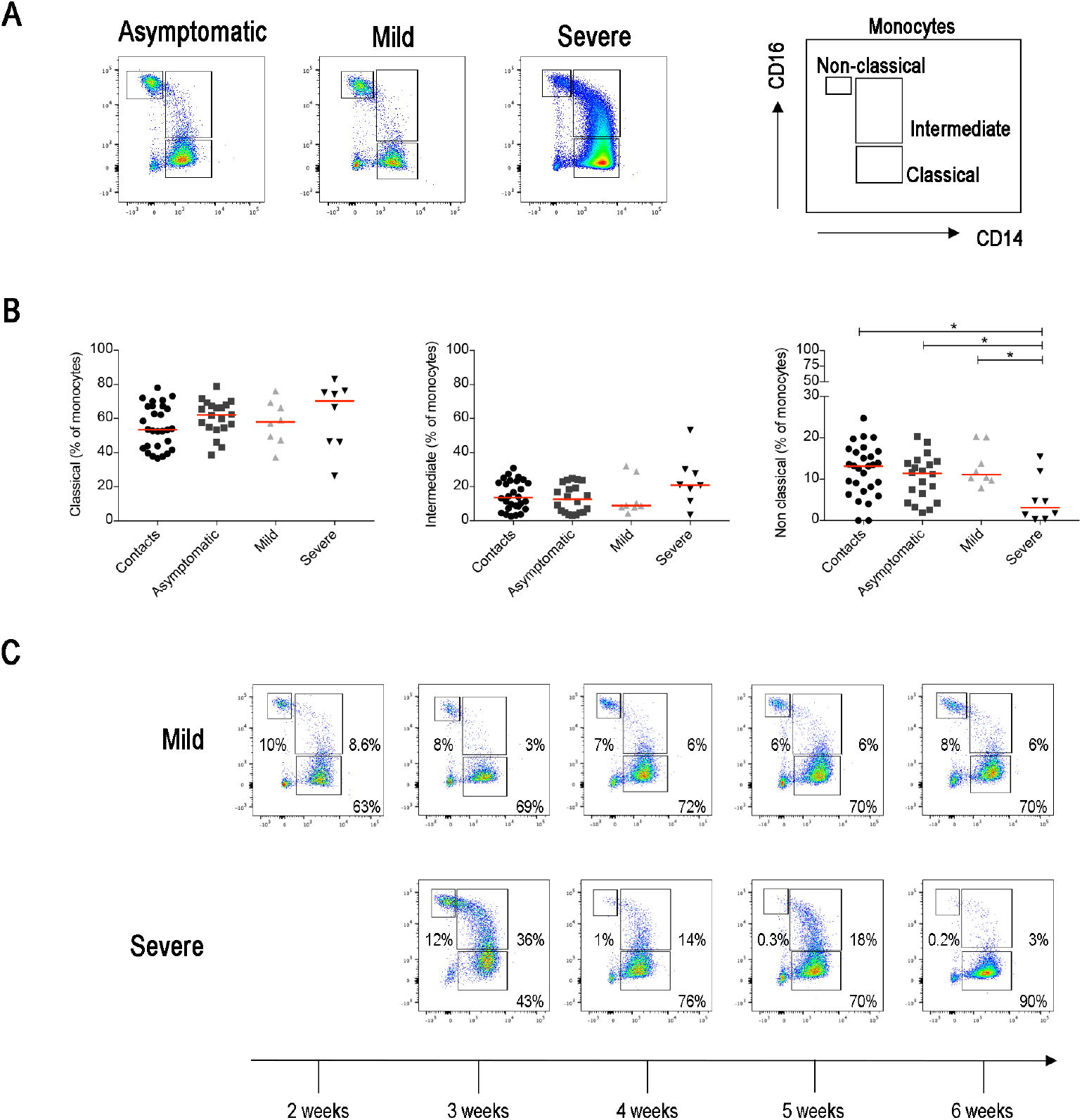
(**A**) Flow-cytometry analysis of monocytes in the blood of three representative patients with asymptomatic, mild and severe disease. Monocytes were gated as CD3^-^ FSC-A^high^. Monocytes were divided as follow: classical (CD14^++^CD16^-^), intermediate (CD14^+^CD16^+^) and non-classical (CD14^+^CD16^++^). (**B**) Scatter plots indicate the percentage of classical, intermediate and non-classical monocytes in each group of patients reported as single value. Midlines indicate median. Statistical significances were determined using unpaired, two-tailed Mann-Whitney *U*-tests. *p≤0.05, **p<0.01, ***p<0.001. (**C**) FACS plots show the different distribution of monocytes populations in two representative patients (one mild and one severe) during the course of the disease (2-6 weeks).

**Supplementary Figure S4:**
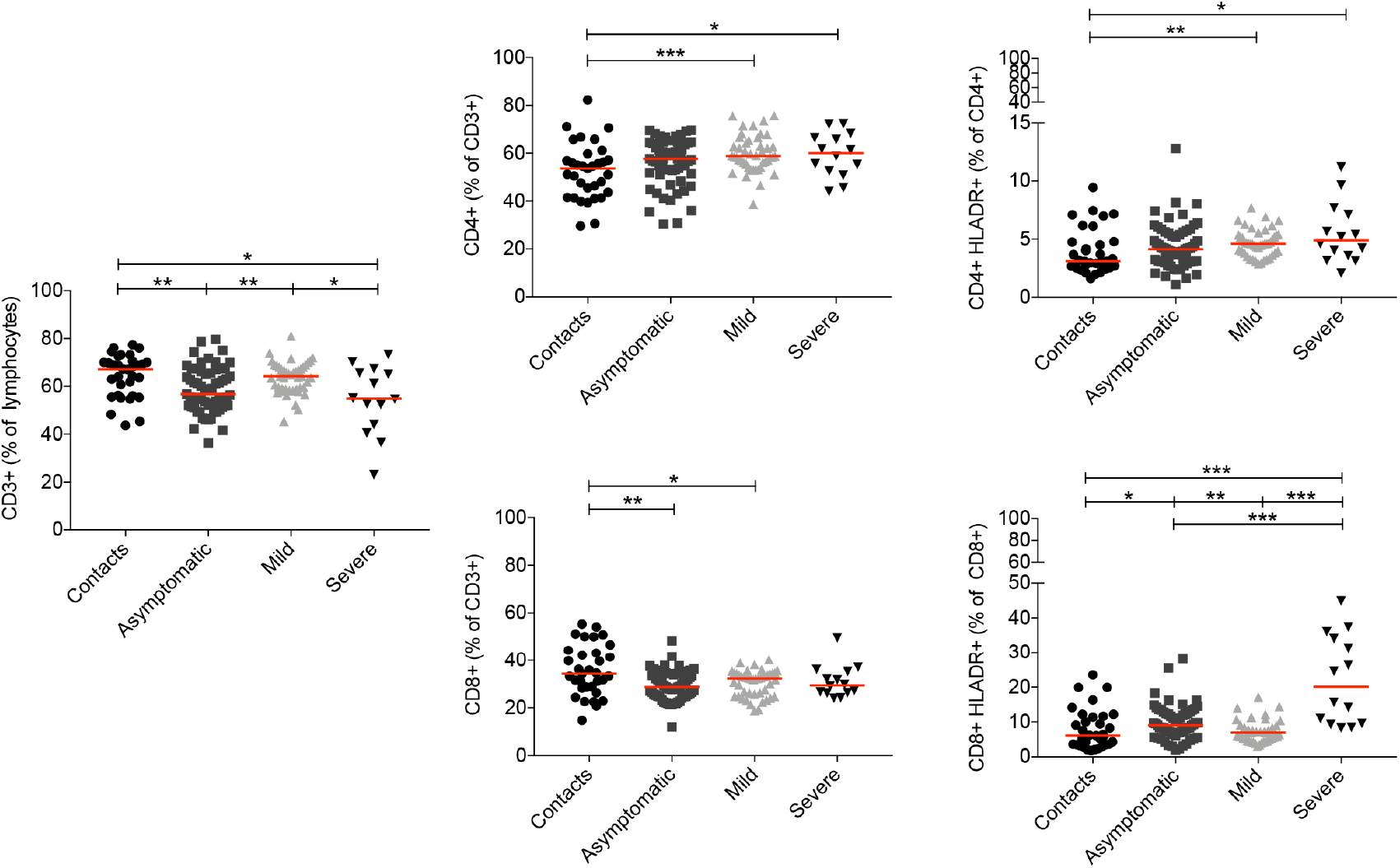
Scatter plots show percentage of T cells (CD3^+^), CD4^+^, CD4+HLA-DR^+^, CD8^+^ and CD8^+^HLA-DR^+^ T cells in all sample analyzed in the study. Midlines indicate median. Statistical significances were determined using unpaired, two-tailed Mann-Whitney *U*-tests. *p≤0.05, **p<0.01, ***p<0.001.

**Supplementary Figure S5:**
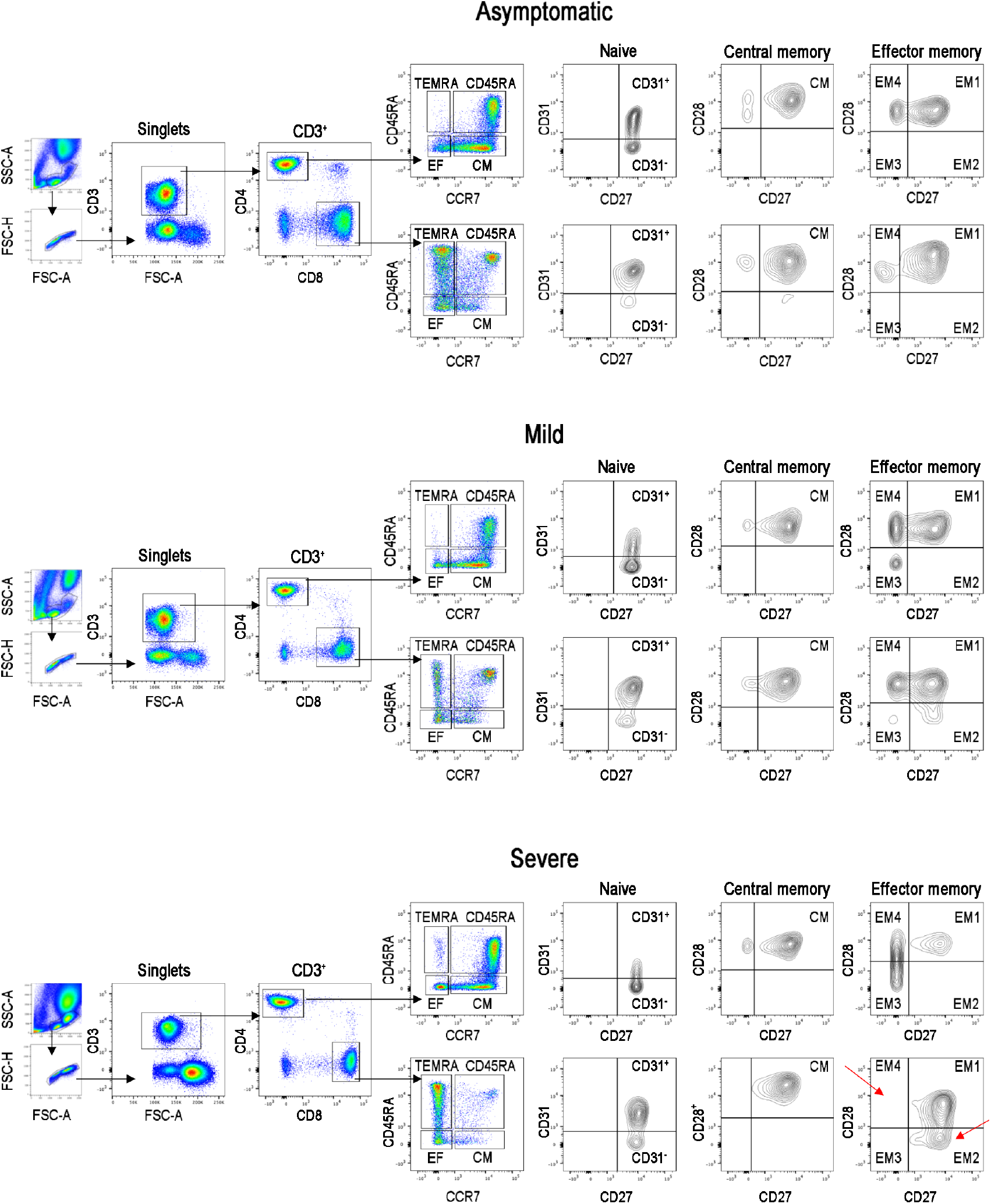
The CD4^+^ and CD8^+^ T cells were subdivided into the main T cell subsets using CD45RA and CCR7: *naïve* (CD45RA^+^CCR7^+^), central memory (CM CD45RA^-^CCR7^+^), effector memory (EM CD45RA^-^CCR7^-^) and TEMRA (CD45RA^+^CCR7^-^) T cells. Naïve T cells were further divided based on CD31 expression (CD31^+^ and CD31^-^). CM and EM were separated based on the expression of CD27 and CD28.

**Supplementary Figure S6:**
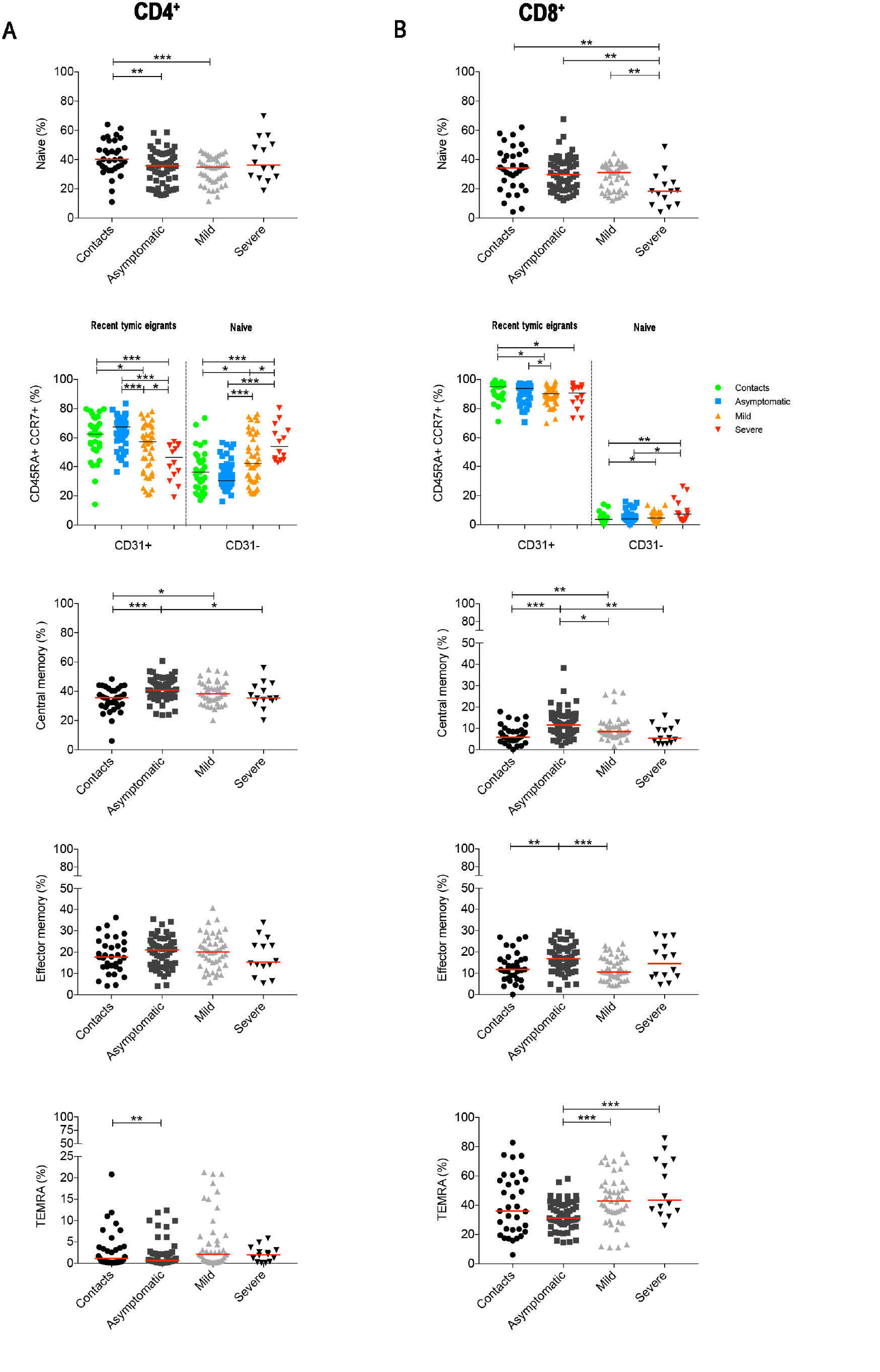
(**A-B**) We separately show the results obtained by the analysis of cumulative data from all samples of CD4^+^ (**A**) and CD8^+^ T cells (**B**). Graphs show the percentage of naïve T cells (CD3^+^CCR7^+^CD45RA^+^) that either expressed or lack CD31 (CD31^+^ and CD31^-^). Percentage of central memory (CD3^+^CCR7^+^CD45RA^-^), effector memory (CD3^+^CCR7^-^CD45RA^-^) and TEMRA (CD3^+^CCR7^-^CD45RA^+^) T cells were shown. Midlines indicate median. Statistical significances were determined using unpaired, two-tailed Mann-Whitney *U*-tests. *p≤0.05, **p<0.01, ***p<0.001.

**Supplementary Figure S7:**
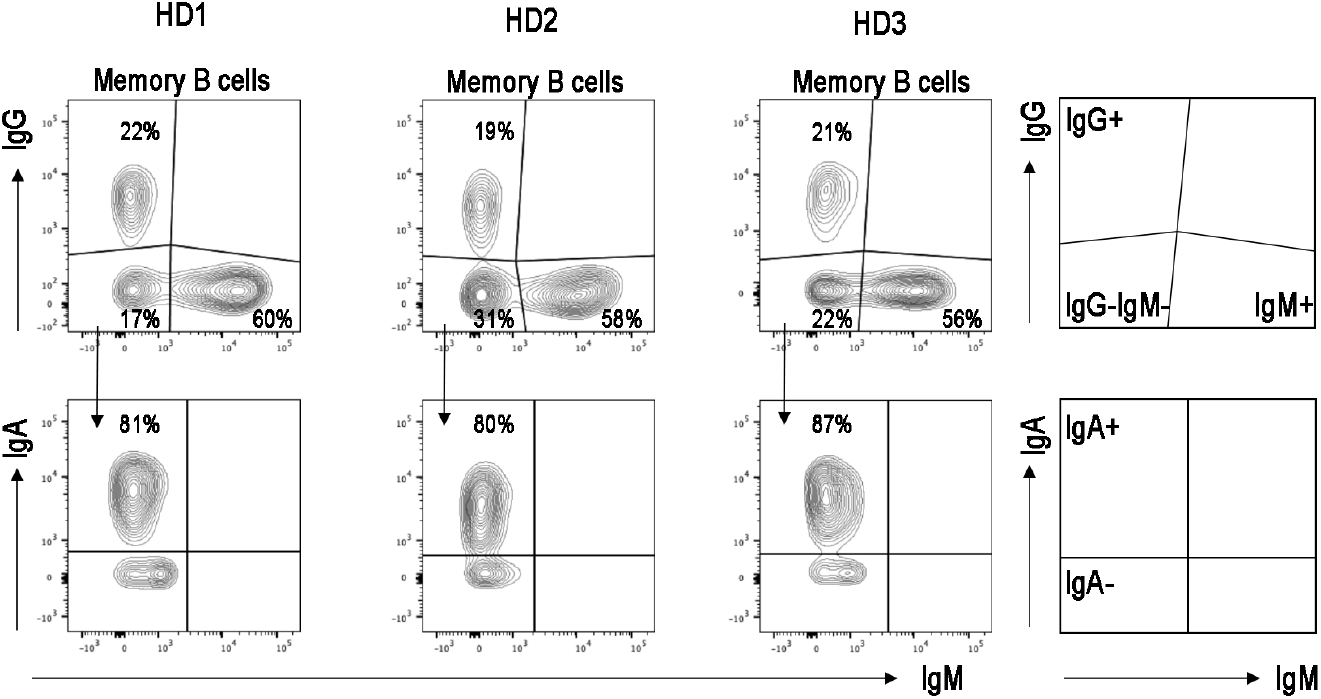
FACS plots in three healthy donors indicates that most of IgG^-^IgM^-^ MBCs correspond to IgA expressing MBCs and minimal part of these cells are IgA^-^IgG^-^IgM^-^ MBCs.

**Supplementary Figure S8:**
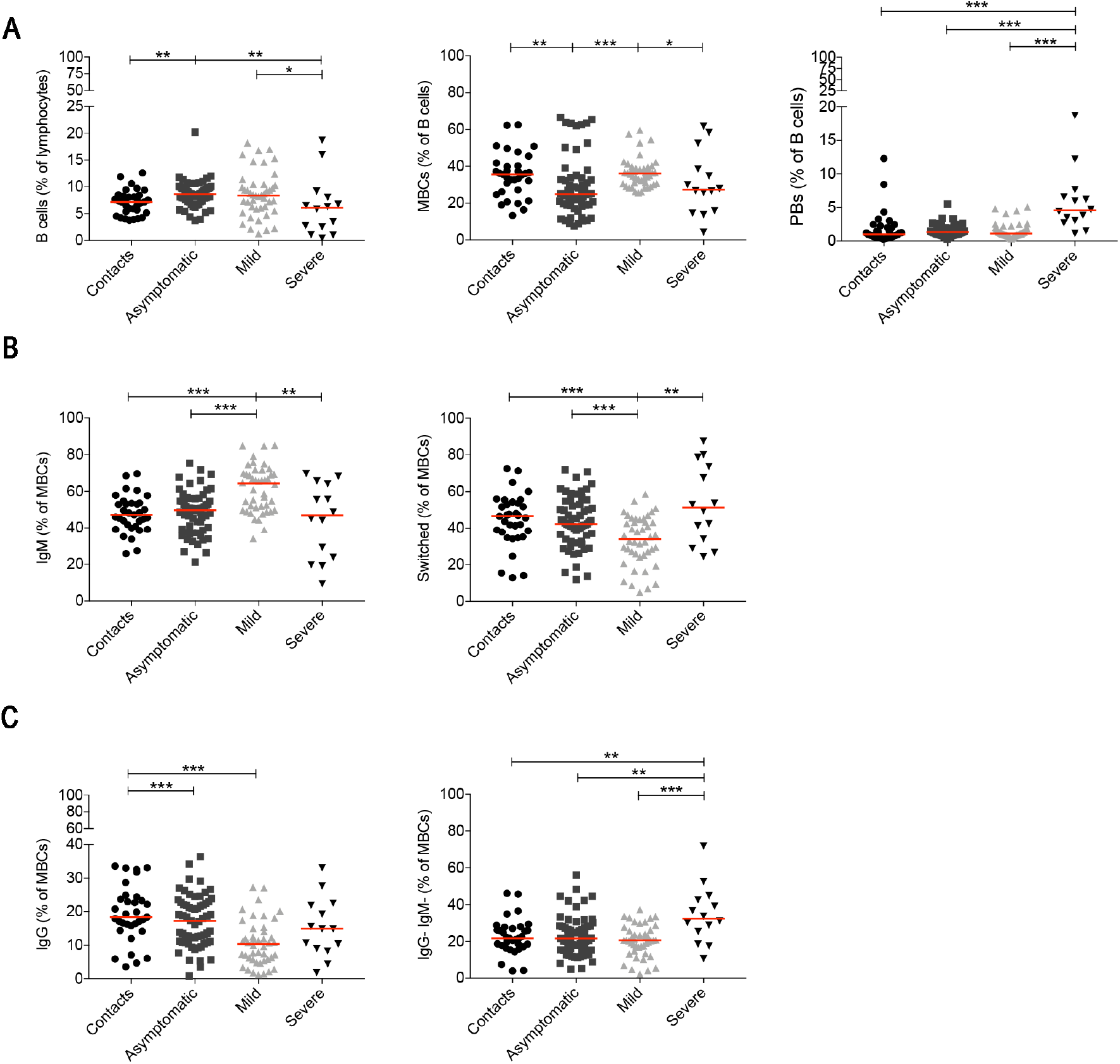
(**A**) Plots indicate the percentage of B cells, MBCs and plasmablasts. In **B**) we show the frequencies of IgM and switched MBCs. In **C** we show the frequency of IgG^+^, IgG^-^ IgM^-^MBCs. (**A-C**) Graphs refer to all samples analyzed in the study. Midlines indicate median. Statistical significances were determined using unpaired, two-tailed Mann-Whitney *U*-tests. *p≤0.05, **p<0.01, ***p<0.001.

